# Passive smartphone-based assessment of cognitive changes in neurosurgery

**DOI:** 10.1101/2020.11.10.20228734

**Authors:** Kevin Akeret, Flavio Vasella, Olivia Zindel-Geisseler, Noemi Dannecker, Peter Brugger, Luca Regli, Peter Young, Martin N. Stienen, Arko Ghosh

**Affiliations:** Department of Neurosurgery, University Hospital Zurich & Clinical Neuroscience Center, University of Zurich, Zurich, Switzerland; Laboratory of Molecular Neuro-Oncology, Clinical Neuroscience Center, Department of Neurology, University Hospital Zurich, Zurich, Switzerland; Neuropsychology Unit, Department of Neurology, University Hospital Zurich, University of Zurich, Zurich, Switzerland; University Hospital of Psychiatry, Zurich, Switzerland and Valens Rehabilitation Centre, Valens, Switzerland; Lancaster Environment Centre and the Data Science Institute, Lancaster University, United Kingdom; Cognitive Psychology Unit, Institute of Psychology, Leiden University, Leiden, The Netherlands

**Keywords:** Neurosurgery, Smartphone behavior, Processing speed, Cognitive testing, Inductive model, Postsurgical recovery

## Abstract

Clinical observations suggest dynamic alterations in behavior after brain surgery. While some alterations reportedly occur within days others gradually develop over several months. These alterations can be attributed to the pre-surgical impact of the diseased tissue, neuronal damage caused by the surgery, and subsequent plasticity. A key step towards a systems-level understanding of the brain-behavior relationships is to capture the dynamics of the cognitive alterations. Here, we first established in 38 healthy individuals that the day-to-day smartphone interactions can be used to inform on cognitive processing speed. Next, we analyzed the smartphone interactions in 12 patients undergoing intracranial tumor surgery, with postsurgical follow-up of up to a year. In healthy individuals, the speed of the touchscreen interactions was highly correlated to choice reaction times (R^2^ = 0.71) but less so to simple reaction times (R^2^ = 0.15) on Deary-Liewald tests. Touchscreen interactions slowed immediately after surgery but the post-surgical changes varied between patients. Data-driven models revealed the time-constant of the short-term postsurgical changes and the time taken to stabilize after the surgery. Furthermore, by using conceptually distinct types of touchscreen interaction speeds – i.e. unlocking time and app locating speed – we established that the post-surgical changes are domain-specific. Interestingly, in this small sample, the pre-surgical smartphone speeds were highly related to the speeds post-stabilization (R^2^ = 0.75 to 0.95). The proxy measures of cognition seamlessly captured on the smartphone can reveal postsurgical dynamics inaccessible to conventional testing. We propose that the transient cognitive alterations indicate the time-constrained influence of distinct neuronal mechanisms triggered by the surgery.

## Introduction

The functional outcome after neurosurgery is difficult to predict. This uncertainty can be attributed to the complex mapping between brain functions and behavior, as well as the diverse ways in which the diseased tissue and its removal influence brain functions (Kiecolt-Glaser et al., 1998). Thus, intact brain areas may help compensate for the brain tissue damaged by the surgery itself (Duffau et al., 2006; Sokolov et al., 2014). Furthermore, the disease itself introduces substantial functional alterations that may already be evident in the presurgical period. For instance, 80% of the patients suffer from some neurocognitive deficits at diagnosis, and these are related to tumor location and elevated intracranial pressure (Habets et al., 2014; Hendrix et al., 2017; Liouta et al., 2016; Satoer et al., 2012; Talacchi et al., 2011). Routine clinical observations suggest that cognitive performance fluctuates after the surgery. An objective assessment of the cognitive fluctuations is a necessary step towards empirical and theoretical development on how the brain responds to invasive perturbation. How quickly does the brain respond to the surgery? To what extent are the alterations transient vs. permanent? Addressing these questions may offer crucial insights into the underlying mechanisms.

Although the post-surgical cognitive dynamics are extremely relevant, patients do only undergo sparse (1-2) follow-up functional assessments in clinical routine. For instance, in brain tumors and other intracranial surgeries, such longitudinal functional assessments play a key role in identifying postsurgical complications (Bartek et al., 2015). However, commonly used functional assessment instruments, such as the Karnofsky performance (KP) scale, only capture relatively severe changes because they rely on subjective reports. Their administration is also time-consuming. For instance, the KP scores are based on the self-reported performance in real-world tasks collected via interviews by the medical staff (Schag et al., 1984). Moreover, as these assessments are often only conducted sparsely, they can mostly inform on the inter-individual differences rather than help resolve the dynamics in a specific patient in the absence of drastic alterations in performance (Meyers & Brown, 2006).

The temporal course of the pre- and post-surgical changes in function may be non-linear and fluctuate considerably over various time-scales. While most patients show some improvement over the weeks following surgery, 20% of the patients deteriorate in the subsequent months (Habets et al., 2014; Talacchi et al., 2011). Neuropsychological assessments of the functional alterations in the days immediately following the surgery are resource-intensive and time-consuming and, more seriously, suffer from individual differences in test-retest susceptibility. Also, they are likely biased by surgical after-effects (e.g. headache, side-effects of anesthesia). Such assessments have, therefore, remained elusive.

The day-to-day digital behavior offers a fresh opportunity to capture the dynamic alterations in neurosurgery. The speed of performance – such as measured in reaction time tasks – is a common quantitative indicator of cognitive status. Interestingly, activities like typing can act as a proxy measure of processing speed (Austin et al., 2011). There is emerging evidence that processing speed can be proxied via the ubiquitous touchscreen smartphone interactions. First, the speed of the day-to-day smartphone interactions shows strong correlations to the extent of trial-to-trial motor variability on tactile reaction time tasks (and a weaker correlation to the reaction time itself)(Balerna & Ghosh, 2018). Second, the speed of interactions shows a strong diurnal rhythm akin to the fluctuations in reaction time performance (Huber & Ghosh, 2020). A straightforward assessment on whether the interaction speed can faithfully proxy the commonly used visual reaction time task is presented here.

In this study, we first established which cognitive processes are captured by the background touchscreen measures. Towards this, we tested the relationship strength between reaction time performances – in a simple and 4-choice task – with that of the touchscreen measures (Deary et al., 2011). We anticipated a relationship between the tests and smartphone behavior, but a priori it was not clear if and how the relationship strength depended on the task used. Next, we deployed the background touchscreen measures on patients scheduled for tumor resection neurosurgery. While we did anticipate behavioral fluctuations, we could not anticipate their patterning, and thus relied on data-driven models to capture and explore the impact of the surgery on the proxy measures of cognitive performance (Young, 1984).

## Methods

### Participants

This was a prospective observational study approved by the local ethical-committee (KEK-ZH BASEC 2018-00395) and registered at www.clinicaltrials.gov (NCT03516162). Neurosurgical patients were assessed for eligibility. The inclusion criteria for the patients comprised: Age over 18, fluent language-skills in German, scheduled for resection of a brain tumor, unshared smartphone use (Android operating system; >3 months), minimum presurgical baseline recording of 7 days, and written informed consent. Reasons for exclusion were any neurologic or psychiatric disease other than a brain tumor, which could potentially influence neurocognition, or foreseeable difficulties in follow-up (e.g. geographic reasons). Twelve patients were recruited with one patient prematurely dropping out and so only yielding pre-surgical measurements. The ages ranged from 31 to 72 years, with a median of 48 years.

Healthy volunteers were extracted from the database raised via agestudy.nl (a Leiden University lifespan study), with the data compiled on the 20^th^ of October 2020. The data collection contains a co-registration of smartphone behavior and online cognitive testing. All participants were recruited online and included self-reported healthy participants. In this collection, we found 38 participants that met the analysis requirements of this study i.e., test administered on a PC and smartphone behavior logs overlapping in a 24 h window (see below). The ages ranged from 21 to 75 years, with a median age of 50 years. This collection was approved by the Psychology Research Ethics Committee at Leiden University.

### Clinical examination for patients

All patients underwent a general functional and neurocognitive examination pre-surgically, within the first week after surgery; and at 3-months-follow-up, including the KPS and the Montreal Cognitive Assessment (MoCA). Clinical and magnetic resonance imaging (MRI) examinations were conducted along the same time-frame to determine tumor location, size, and extent of resection. The patients self-reported their handedness. On recruitment, patients installed the TapCounter App (QuantActions, Lausanne) (Balerna & Ghosh, 2018). The T-scores of MoCA were estimated based on normative data available in the literature (Thomann et al., 2018).

### Smartphone data recording

Smartphone touchscreen interactions were recorded using the TapCounter App (QuantActions LTD, Lausanne) (Balerna & Ghosh, 2018). In brief, the touchscreen timestamps were recorded along with the corresponding App labels. The App operated in the background and the data were downloaded via taps.ai (QuantActions LTD). Relevant users on taps.ai were identified using alphanumeric identifiers. The downloaded data were parsed into MATLAB using custom-written scripts (available on Taps.ai). The additional permission necessary for App labels meant that the corresponding metrics were not available in one healthy participant.

### Smartphone data analysis

We extracted three proxy measures of processing speed: (i) Tapping speed (TS): Shortest 25^th^ percentile inter-touch intervals from each session (between screen ON and OFF) collected in 24 h bins. The median of these measures was used to describe the TS in 24 h bin. (ii) Unlocking speed (US): Shortest 25^th^ percentile inter-event intervals in 24 h bins, between the screen turning ON and the first touch recorded on the unlocked screen. (iii) App locating speed (ALS): Shortest 25^th^ percentile inter-touch intervals in 24 h bins, occurring between two consecutive touches on the home or launch screen before an app touch registration. In the patient population, the 24 h bin onset edges were at 00:01, whereas in the healthy population a 24 h window encompassing the PC-based reaction time testing was used (so if the test was conducted at 09:00, the bin edges were at 21:01 of the previous day and 21:00 of the same day).

### The Deary-Liewald reaction time task

The Deary-Liewald visual reaction time tasks – simple and 4 choice – were implemented on agestudy.nl running Psytoolkit (Deary et al., 2011; Stoet, 2010, 2017). In brief, a test session contained 50 test trials each for the simple and choice tasks. These test trials were preceded by 8 trials for adjusting to the task. The task order was set to simple and then choice. The right hand (thumb on space bar) was used for the simple task. For the choice task, the index and middle fingers of both hands were in use mapped to the 4 locations on the screen. The test was administered either in Dutch or English based on the language preference of the user. The code used towards the English version of the test is shared on Psytoolkit.

### Data-driven models and statistical tests

The impulse response functions were fitted using the CAPTAIN (Taylor et al., 2007; Young, 1984) toolbox on MATLAB (MathWorks, Natick) and the parameters were selected using the coefficient of determination based on the model output 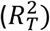. A robust (bi-square) fitting method was used in the linear regression analysis (*fitlm* function on MATLAB). The MATLAB codes used this study are available at the repository link.

## Results

### Smartphone-based metrics collected in the background reflect choice reaction time performance

We compared the reaction time performance to the background smartphone behavioural metrics aimed as a proxy measure of cognitive processing speed (**Fig. 1**). Not surprisingly, the choice reaction times were consistently longer than the simple reaction times (simple Mean = 301 ms; choice Mean = 511 ms; *p* < 0.001; *t*(59) = −13.2, paired t-test). We detected weak but significant correlations between the simple reaction time vs. some of the smartphone metrics (vs. Tapping speed [TS], R^2^ = 0.147, β = 52.7, *t*(36) = 2.44, *p* = 0.018, bi-square linear regression fit; vs. Unlocking speed [US], R^2^ = 0.003, β = −5.23, *t*(36) = −0.24, *p* = 0.812; vs. App locating speed [ALS], R^2^ = 0.112, t(35) = 2.09, *p* =0.044). The correlations between the choice reaction time vs. the smartphone metrics were strong (vs. TS, R^2^ = 0.71, β = 277.19, *t*(36) = 9.28, *p* = 4.39 x 10^−11^; vs. US, R^2^ = 0.014, β = 38.507, *t*(36) = 0.72, *p* = 0.48; vs. ALS, R^2^ = 0.50, β = 392.03, *t*(35) = 5.88, *p* = 1.13 x 10^−06^). Given the strong correlations for TS and ALS, we next addressed their separate contribution in explaining the variance in choice reaction time. The multivariate model between the reaction time vs. smartphone metrics revealed a stronger relationship with TS than ALS (R^2^ = 0.73, *f*(33) = 29.6, *p* = 1.77 x 10^−9^, linear multi-regression model; β _*TS*_ = 194.52, *t* = 4.99, *p* = 1.92 x 10^−05^; β_*US*_ = −18.9, *t* = −0.68, *p* = 0.5; β_*ALS*_ = 197.63, *t* = 3.05, *p* = 0.0045). In sum, smartphone-based metrics captured in the background reflect the reaction time performance with each metric capturing different aspects of the cognitive test.

**Figure 1.**
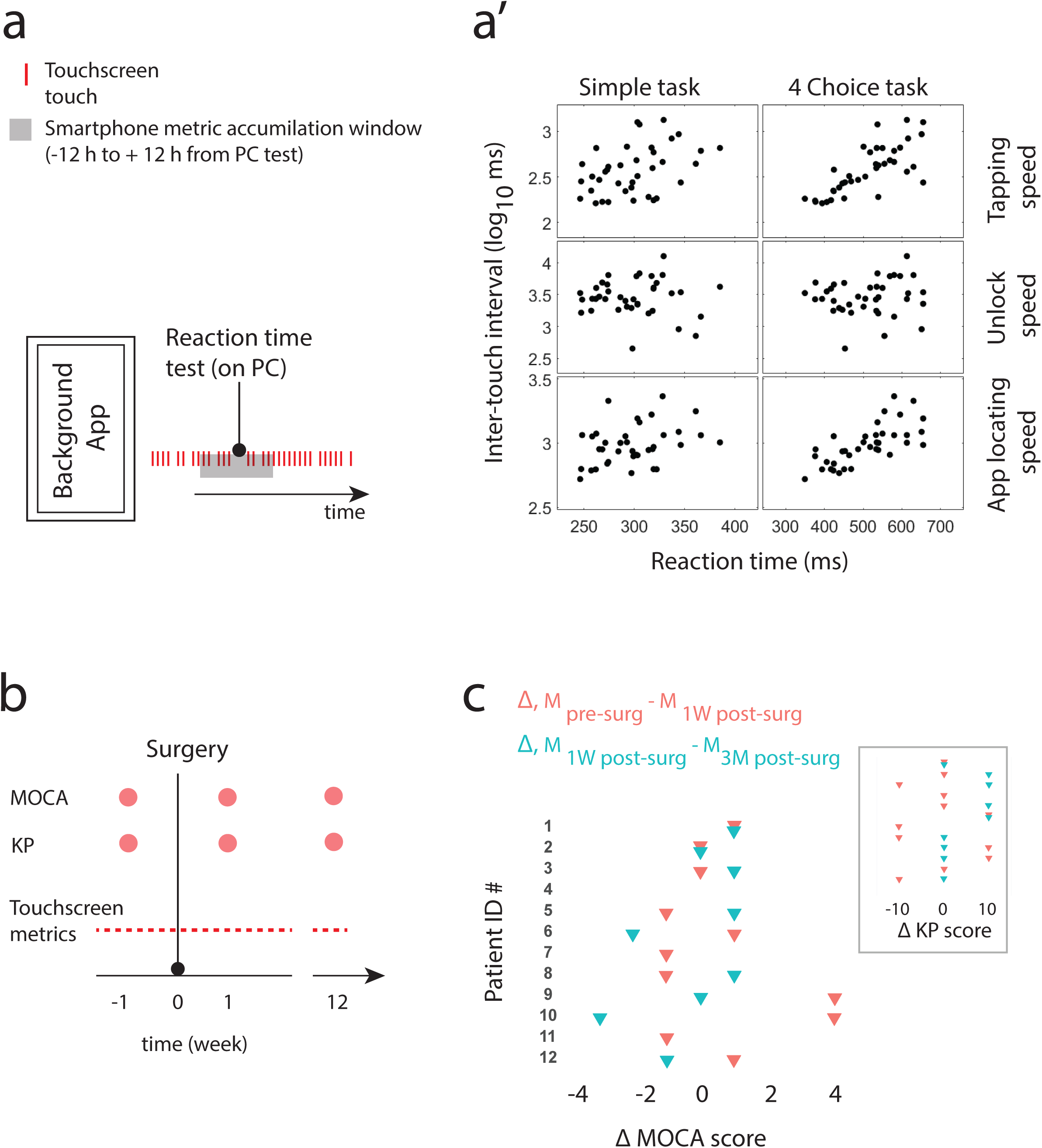
Smartphone behaviour can proxy reaction time performance and standard clinical assessments of cognitive status in neurosurgical patients. (a’) A schematic diagram of the observational set-up used to link the smartphone parameters and reaction time performance measured on a PC. (a) Correlation matrix between reaction time performance the smartphone parameters gathered on the day of the reaction time test. (b) Standard clinical assessments – MoCA and KP performance scores – were obtained at three different time points. (c) Change in Montreal Cognitive Assessment (MoCA) and Karnofsky performance scale (KP, insert) scores before and after the surgery.

### Overview of patients

Key patient and tumour characteristics are described in **Table 1**. Patients were monitored pre-surgically for 21 d (median, range: 6 to 38 d) and 144 d (median, range: 83 to 388 d) post-surgically. Pre-surgical smartphone usage was related to the age of the patient but not the tumour diameter (n = 12, *R*_2_ = 0.52, *f*(9) = 4.8, *p* = 0.038, linear multi-regression model; β_*age*_ = −0.033, *t* = −3.06, *p* = 0.014; β_*tumour size*_ = 0.016, *t* = 1.21, *p* = 0.26). All of the users were found to be back on the device within 3 days post-surgery.

**Table 1.**
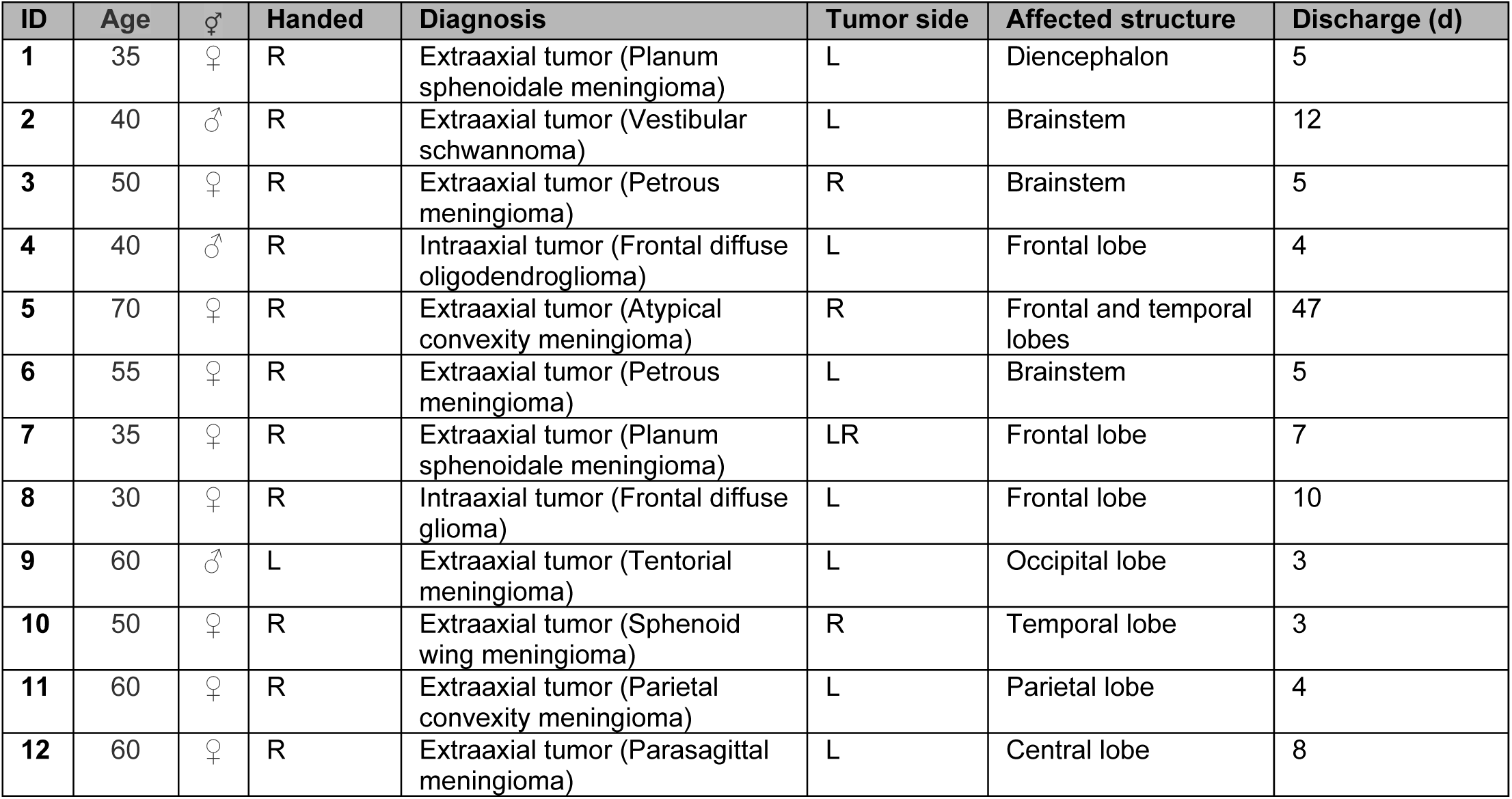
Key patient and tumour characteristics. The same patient IDs are as used in the figures and in the text. L: Left, R: Right. The postoperative day (d) of hospital discharge is reported in the ‘Discharge’ column. Ages are rounded to 5 years.

### MoCA performance in patients

There was no consistent pattern of variance introduced by the surgery in the MoCA scores (**Fig. 1**). Interestingly, the MoCA scores collected at 1 week after surgery were no different from the normative healthy data (T scores, 45.5; 56.2; 50.6; 61.9; 60.5; 62.7; 47.8; 24.7; 49.3; 53.7; 48.7; 47.2). The KPS showed even less variance than the MoCA scores and were not considered further for in-depth analysis. The presurgical TS, in terms of inter-touch interval, revealed an anticipated relationship with age, such that the interval was larger in older individuals (n = 12, *R*_2_ = 0.67, *f*(9) = 9.02, *p* = 0.007, linear multi-regression model, β_*age*_ = 0.013, *t* = 3.80, *p* = 0.004). The other presurgical inter-individual differences in processing speed – US and ALS – showed no relationship to age or tumor diameter. Pooled MoCA measures were related to the processing speed (TS) captured on the smartphone on the corresponding days; the larger the MoCA score the shorter the inter-touch interval (*R*_2_ = 0.30, β = −0.064, *t*(30) = −3.53, *p* = 0.001).

### Behavioral dynamics after the surgery

Concerning the continuously measured smartphone parameters in the patients, we anticipated that fluctuations after the surgery are not simply linear and, therefore, used an inductive modeling approach to capture the surgical after-effects in the form of transient impulse response functions. This allowed for the construction of model systems that can be systematically explored and attached to this report. Here we describe the key features of the model. The models captured the diverse behavioral patterns, time-locked to the surgery, for the three different smartphone parameters (**Fig. 2**). Interestingly, the captured patterns varied both across the patients and the parameter used to capture processing speed. The response functions used to model the continuous measures as impulse responses (with the surgery time modelled as an impulse) revealed that some instances were dominated by rapid processes governed by a time-constant of 7 days (median, Q1 1.5 d – Q3 46.5 d). The period for the parameters to stabilize (proxied by using the settling time of the function) was 51.5 days (median, Q1 14.7 d – Q3 136.2 d). Notably, the slower the time-constant underlying the rapid behavioural change immediately after the surgery, the more delayed was the stabilization in the long-term (*R*_2_ = 0.23, β = 0.347, *t*(22) = 2.57, *p* = 0.017, linear regression, pooled parameters from fitted models of *R*_2_ > 0.15). We finally addressed whether pre-surgical behavior was correlated to the performance in the final seven days of the observation (i.e., post stabilization). The values were highly correlated for all of the smartphone parameters (TS, *R*_2_ = 0.85, β = 0.87, *t*(9) = 7.01, *p* = 6.26 x 10^−5^; US, *R*_2_ = 0.93, β = 1.15, *t*(9) = 10.74, *p* = 1.97 x 10^−6^; ALS, *R*_2_ = 0.88, β = 0.81, *t*(9) = 8.27, *p* = 1.70 x 10^−5^).

**Figure 2.**
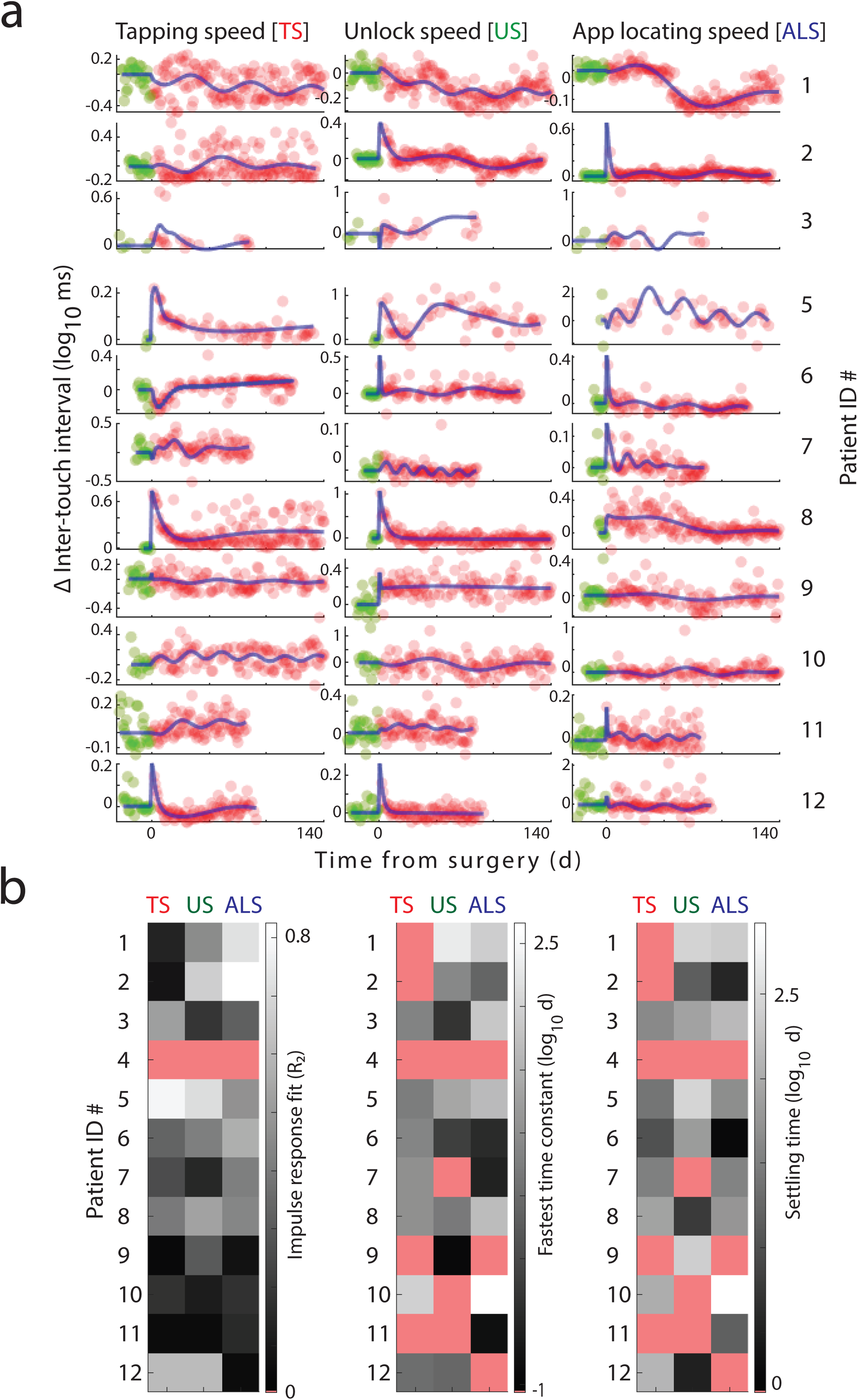
Dynamic post-surgical alterations in cognitive speed captured on the smartphone. (a) The pre- (in green) and postsurgical (in red) smartphone parameters captured in each patient. Transient impulse response model fitting (blue solid lines) used all of the data points in each patient (the display is trimmed to 140 days for visualisation). (b) The R2 values of the impulse response fit for all the patients and parameters. Note, patient ID #4 did not have postsurgical data (marked in pink). The time-constants underlying the fastest postsurgical fluctuations and the time to stabilize (impulse response settling times) are reported for only for fits R2 > 0.15 (under threshold values are marked in pink).

## Discussion

We leveraged smartphone interactions to quantify the dynamic behavioral fluctuations surrounding intracranial tumor resection. The dense quantification revealed a highly personalized account of behavioral fluctuations, where changes can be observed immediately after the surgery but may take months to stabilize. These dynamic fluctuations could not be traced using standard clinical assessments such as KPS and MoCA scores and offers unique perspectives on the mechanisms underlying spontaneous recovery.

Our observations in healthy individuals show that the cognitive processes as captured in reaction time assessments reflect in smartphone behavior. The finding that visual simple reaction time is weakly correlated to smartphone tapping speed (TS) is in line with our previous observation on tactile reaction time (Balerna & Ghosh, 2018). In our previous study, we observed that motor variability was strongly related to TS raising the possibility that this parameter captures more complex cognitive processes than input-output operations. This was confirmed here with the finding that visual choice reaction time is strongly correlated to TS. In sum, the smartphone-based parameters may be used to proxy higher sensorimotor processes rather than simple sensorimotor computations. Moreover, according to our observation in the patients, the MoCA scores were moderately correlated to TS, suggesting that smartphone behavior is loosely linked to the general cognitive status.

Neither the KPS nor the MoCA scores showed any clear fluctuations induced by the surgery. This is perhaps not surprising considering that carefully conducted microneurosurgery must not necessarily lead to measurable alterations of brain function – at least using these conventional measures of cognitive health status. Notably, in comparison to a healthy database, the MoCA scores did not offer any decisive information. These observations suggest that although MoCA and KPS scores may be sensitive to large cognitive alterations as induced by substantial brain injury, they are limited in their ability to construct individual behavioral dynamics in less invasive neurosurgery as studied here.

A diversity of fluctuations was captured in the smartphone parameters surrounding the surgery. In all but two patients we observed immediate changes in the proxy cognitive measures. These initial alterations – governed by a time constant of ∼ 7 days – may be driven by mechanical perturbations of brain activity by edema or rapid neuronal adaptations involving excitation-inhibition modulations. There was substantial diversity in the time constant of these fluctuations indicating that the underlying mechanisms operate differently across individuals and cognitive processes. Patients #1 and #10 showed no rapid behavioral fluctuations. Notably, both of these patients had smaller tumors and lower smartphone usage than the rest of the sample. Nevertheless, patient #1 showed a substantial improvement in one of the parameters two months after the surgery.

In addition to the rapid smartphone parameter fluctuations, we also captured slower changes with a time constant of ∼ 50 days. These slower fluctuations may result from gradual forms of neuronal plasticity for instance via use-dependent structural alterations. Interestingly, the time taken for the parameters to stabilize were strongly related to the time taken for the initial fluctuations. This raises two distinct possibilities. One, that the late changes in cognition are triggered by the early mechanisms. Two, that surgical features – such as the extent of the mechanical perturbation - dictate both the early and late alterations. Perhaps a combination of the two is as likely, and further exploration is needed where the background measures are combined with longitudinal neuroimaging to help resolve this.

An important question here is, what determines the long-term behavioral outcomes (after stabilization of the post-surgical behavioral dynamics)? One possibility is that the factors such as the location of the surgery or extent of tissue damage are the main determinants. Another possibility is that the long-term consequences are determined by the pre-surgical state. In our limited sample, we found a strong relationship between the pre-surgical smartphone parameters and post-stabilization performances. Essentially, people largely return to their pre-surgical performance levels rather than obtaining an entirely different level of performance. This raises the interesting possibility that despite the turbulent behavioral dynamics stable attractors are guiding behavioral outputs in the long-term.

The fast and the putative, slow responses were not simply linear, which indicates that the sparse measurements employed in routine clinical practice may lack the necessary temporal resolution to adequately capture postsurgical outcomes (Meyers & Brown, 2006). While the clinical relevance of these fluctuations remains to be established, it is conceivable that the identification of specific patterns could lead to improved patient care, be it through early recognition of complications, or evaluation of rehabilitation progress. The inter-individual differences in the behavioral patterns may be explained by the various locations of the lesions and their corresponding neurocognitive function, resulting in different surgical approaches, patient age, post-surgical treatments, pre-surgical state, and the nature of the tumor itself (Liouta et al., 2016). Addressing these parameters is a logical next step.

Taken together, we demonstrate that smartphone interactions, collected from everyday usage, can be utilized to monitor the cognitive status in the perioperative setting of brain tumor surgery. Not only did all the patients quickly return to their smartphones after the surgery but the continued usage allowed for near-continuous monitoring. Moreover, the objective, telemetric monitoring is unobtrusive and inexpensive. This study may serve as a proof-of-concept towards leveraging smartphones as medical sensors in neurology, to complement conventional testing of functional and neurocognitive function. Importantly, the new technology used here raises fresh lines of interrogation on how the brain adapts to the abrupt surgical intervention.

## Data Availability

All data will be made available via a repository link upon peer-reviewed publication.

## Author Contributions

AG and MS conceived the study. KA, FV and MS recruited the subjects and gathered clinical and neuroimaging data. OZG, ND and PB gathered the neuropsychological data. AG analysed the data aided by PY. AG drafted the figures aided by KA. KA, FV, MS and AG drafted the manuscript text. LR, OZG, NG, PB and PY edited the manuscript text.

